# Using past and current data to estimate potential crisis service use in mental healthcare after the COVID-19 lockdown: South London and Maudsley data

**DOI:** 10.1101/2020.06.29.20142448

**Authors:** Robert Stewart, Matthew Broadbent

## Abstract

The lockdown policy response to the COVID-19 pandemic in the UK has a potentially important impact on provision of mental healthcare with uncertain consequences over the 12 months ahead. Past activity may provide a means to predict future demand. Taking advantage of the Clinical Record Interactive Search (CRIS) data resource at the South London and Maudsley NHS Trust (SLaM; a large mental health service provider for 1.2m residents in south London), we carried out a range of descriptive analyses to inform the Trust on patient groups who might be most likely to require inpatient and home treatment team (HTT) crisis care. We considered the 12 months following UK COVID-19 lockdown policy on 16^th^ March, drawing on comparable findings from previous years, and quantified levels of change in service delivery to those most likely to receive crisis care. For 12-month crisis days from 16^th^ March in 2015-19, we found that most (over 80%) were accounted for by inpatient care (rather than HTT), most (around 75%) were used by patients who were current or recent Trust patients at the commencement of follow-up, and highest numbers were used by patients with a previously recorded schizophreniform disorder diagnosis. For current/recent patients on 16^th^ March there had been substantial reductions in use of inpatient care in the following 31 days in 2020, more than previous years; changes in total non-inpatient contact numbers did not differ in 2020 compared to previous years, although there had been a marked switch from face-to-face to virtual contacts.

## Background

The COVID-19 pandemic may have a profound impact on health services through the direct effects of the virus itself, through social distancing policies and their psychological impact, and through the disrupting impact of social distancing policies on delivery of mental and physical healthcare. Mental healthcare services have had to be radically reconfigured to cope with suspected or confirmed COVID-19 infections in inpatient and outpatient settings, staff sickness or self-isolation, the need to minimise face-to-face contacts, and the need to accommodate increasing pressures on acute medical care from cases of viral pneumonia.

We have previously begun reporting on the mental healthcare impact of the UK COVID-19 pandemic, taking advantage of the Clinical Record Interactive Search (CRIS) data platform that receives 24-hourly updates from its source electronic records. Analysing activity in key services from February to mid-May 2020, and comparing periods before and after 16^th^ March 2020, Community Mental Health Teams showed relatively stable caseloads and total contact numbers, but a substantial shift from face-to-face to virtual contacts, while Home Treatment Teams showed the same changeover but reductions in caseloads and total contacts (1).

## Methods

The Biomedical Research Centre (BRC) Case Register at the South London and Maudsley NHS Foundation Trust (SLaM) has been described previously (2;3). SLaM serves a geographic catchment of four south London boroughs (Croydon, Lambeth, Lewisham, Southwark) with a population of around 1.2 million residents, and has used a fully electronic health record (EHR) across all its services since 2006. SLaM’s BRC Case Register was set up in 2008, providing researcher access to de-identified data from SLaM’s EHR via the Clinical Record Interactive Search (CRIS) platform and within a robust security model and governance framework (4). CRIS has been extensively developed over the last 10 years with a range of external data linkages and natural language processing resources (3). Of relevance to the work presented here, CRIS is updated from SLaM’s EHR every 24 hours and thus provides relatively ‘real-time’ data. SLaM’s EHR is itself immediately updated every time an entry is made, which include date-stamped fields indicating patient contacts (‘events’) and those indicating acceptance of a referral or a discharge from a given service (or SLaM care more generally). CRIS data have been substantially enhanced by a suite of natural language processing (NLP) algorithms to extract structured data on named entities from free text fields; a catalogue describing these is available on the CRIS site (5). CRIS has supported over 200 peer reviewed publications to date, and has received approval as a data source for secondary analyses (Oxford Research Ethics Committee C, reference 18/SC/0372).

In order to investigate the composition of service user profiles associated with crisis service use over the 12 months following 16^th^ March 2020 based on previous years’ data, for each recent year (2015-2019), we took the 16^th^ March as an index date and characterised patients who received any crisis care (inpatient or HTT) in the 12 months following each date (i.e. up to and including 15^th^ March in the following year). To begin with, we divided these patients into three cohorts based on their status at the start of follow-up: i) Recent – those active to SLaM at any point in the 3 months up to 16^th^ March that year; ii) Past – those previously active to SLaM but not within the 3 months up to 16^th^ March that year; iii) New – those not previously known to SLaM before the 16^th^ March that year. ‘Active’ here was defined on the basis of an open and accepted referral (i.e. regardless of whether a team had been assigned or a contact made). For each follow-up period, we investigated the proportion of ‘crisis care’ accounted for by inpatient vs. HTT-only. We compared total crisis person-days, inpatient person-days, and HTT-only person-days between the three cohorts for each year.

Further exploratory descriptive analyses of group (i) above divided the 12-month follow-up period into quarters to investigate whether crisis service use was higher in earlier compared to later periods after each index date. Finally, contributions to crisis days over each 12-month period for group (i) were described specifically for seven diagnostic groups, defined on the basis of any previous primary or secondary diagnosis received prior to each index data (thus overlapping rather than mutually exclusive): a) serious mental illness (ICD-10 codes F2x, F30x, F31x, F32.3, F33.3); b) schizophrenia-like disorders (F2x); c) bipolar disorder (F31x); d) any affective disorder (F3x); e) any organic disorder (F0x); f) any alcohol or substance use disorder (F1x); g) any personality disorder (F60x, F61x, or ascertained via an NLP algorithm extracting diagnostic statements in documentation).

In order to quantify and contextualise the recent changes in service contact experienced by current/recent service users in the early stages of lockdown, for each previous year we ascertained patients on March 16^th^ who had been ‘active’ to SLaM in the preceding 3 calendar months (i.e. Group (i) above). Taking these cohorts, we ascertained total inpatient days and the following clinical contacts by non-inpatient teams: i) face-to-face contacts attended; ii) virtual contacts attended (by email, fax, mail, phone, online, or video link); iii) total contacts, as the sum of these two. These were quantified and compared for the 31 days up to and including 15^th^ March, and for the 31 days from 16^th^ March onward, for the years 2015 to 2020. Further analyses were then carried out restricting the comparisons to those with diagnoses recorded prior to 16^th^ March in a given year, applying the same seven diagnostic groups as described above.

## Results

Total crisis days for given 12-month periods are displayed and compared in Table 1, and were around the 350,000 to 360,000 range, tending to be lower in more recent years. Around 80-85% of crisis days were accounted for by inpatient days, reducing gradually from 2015-19. Around 75% of crisis days were used by current/recent patients at the start of each follow-up period (reducing from 77% to 73% from 2015-19); this proportion was lower for HTT days, with around 50-55% being used by recent patients (reducing over the years) and the remainder split fairly evenly between past and new patients.

**Table 1.** Crisis days attributable to current/recent, past, and new mental health service users at the commencement of the 12-month follow-up period

| Year | Cohort | N | Service use in the 12 months from 16<sup>th</sup> March (person-days) |  |  |  |  |  |
| --- | --- | --- | --- | --- | --- | --- | --- | --- |
|  |  |  | All crisis days<br>(inpatient plus HTT) |  | Inpatient days |  | HTT days* |  |
| **2019** | Recent | 3105 | 254377 | 72.8% | 220908 | 78.5% | 36706 | 50.4% |
|  | Past | 1035 | 44124 | 12.6% | 27422 | 9.7% | 17780 | 24.4% |
|  | New | 1403 | 50780 | 14.5% | 33242 | 11.8% | 18367 | 25.2% |
|  | **Total** |  | **349281** |  | **281572** | **80.6%** | **72853** | **20.9%** |
| **2018** | Recent | 3054 | 260593 | 73.8% | 226798 | 79.1% | 36848 | 51.9% |
|  | Past | 992 | 39793 | 11.3% | 23866 | 8.3% | 16655 | 23.5% |
|  | New | 1360 | 52875 | 15.0% | 36225 | 12.6% | 17449 | 24.6% |
|  | **Total** |  | **353261** |  | **286889** | **81.2%** | **70952** | **20.1%** |
| **2017** | Recent | 3074 | 262578 | 75.3% | 230060 | 80.4% | 36139 | 53.3% |
|  | Past | 921 | 37720 | 10.8% | 23528 | 8.2% | 15201 | 22.4% |
|  | New | 1280 | 48198 | 13.8% | 32487 | 11.4% | 16426 | 24.2% |
|  | **Total** |  | **348496** |  | **286075** | **82.1%** | **67766** | **19.4%** |
| **2016** | Recent | 3102 | 276472 | 76.1% | 241020 | 80.5% | 40137 | 57.1% |
|  | Past | 823 | 34641 | 9.5% | 22198 | 7.4% | 13309 | 18.9% |
|  | New | 1246 | 52127 | 14.4% | 36198 | 12.1% | 16878 | 24.0% |
|  | **Total** |  | **363240** |  | **299416** | **82.4%** | **70324** | **19.4%** |
| **2015** | Recent | 3085 | 277800 | 76.6% | 248261 | 80.9% | 33455 | 54.7% |
|  | Past | 865 | 36138 | 10.0% | 24108 | 7.9% | 12810 | 20.9% |
|  | New | 1214 | 48726 | 13.4% | 34557 | 11.3% | 14917 | 24.4% |
|  | **Total** |  | **362664** |  | **306926** | **84.6%** | **61182** | **16.9%** |

HTT: home treatment team
\*Excluding concurrent inpatient and HTT days

In further analyses focusing on current/recent patients at each index date, dividing the 12-month follow-up period into quarters, crisis person-days were observed to be relatively stable over that period for current/recent patients (Figures 1a-c). Descriptions of crisis day usage by diagnostic groups of interest are displayed in Figures 2a and 2b. In summary, highest numbers of crisis days were accounted for by people with previously recorded schizophreniform diagnoses; use of crisis services had been decreasing in SMI, schizophreniform, bipolar and organic disorder diagnostic groups, had been relatively stable in patients with previously recorded affective disorder or alcohol/substance use disorder, and had been increasing in those with a previous personality disorder diagnosis. The proportion of crisis days accounted for by inpatient care (as opposed to HTT) was fairly similar between diagnostic groups, although marginally lower for those with previous affective disorder diagnoses; also, no substantial or consistent changes over the 5 years were observed.

**Figure 1.**
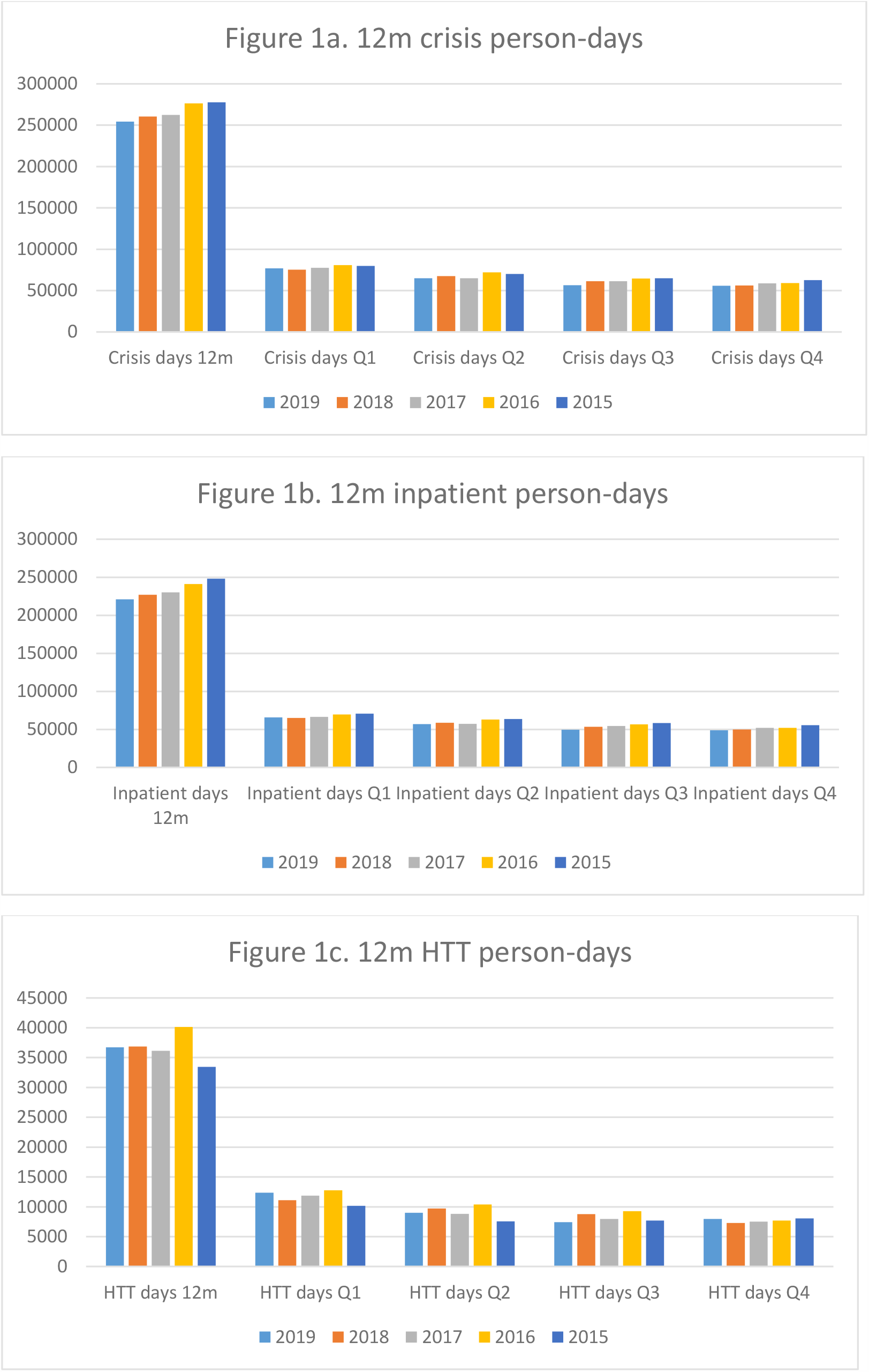
Crisis (home treatment team [HTT] and inpatient care) person-days receipt over 12 month period, for patients with current/recent mental healthcare at the commencement of that period, displaying total days and days by quarters of the follow-up period.

**Figure 2.**
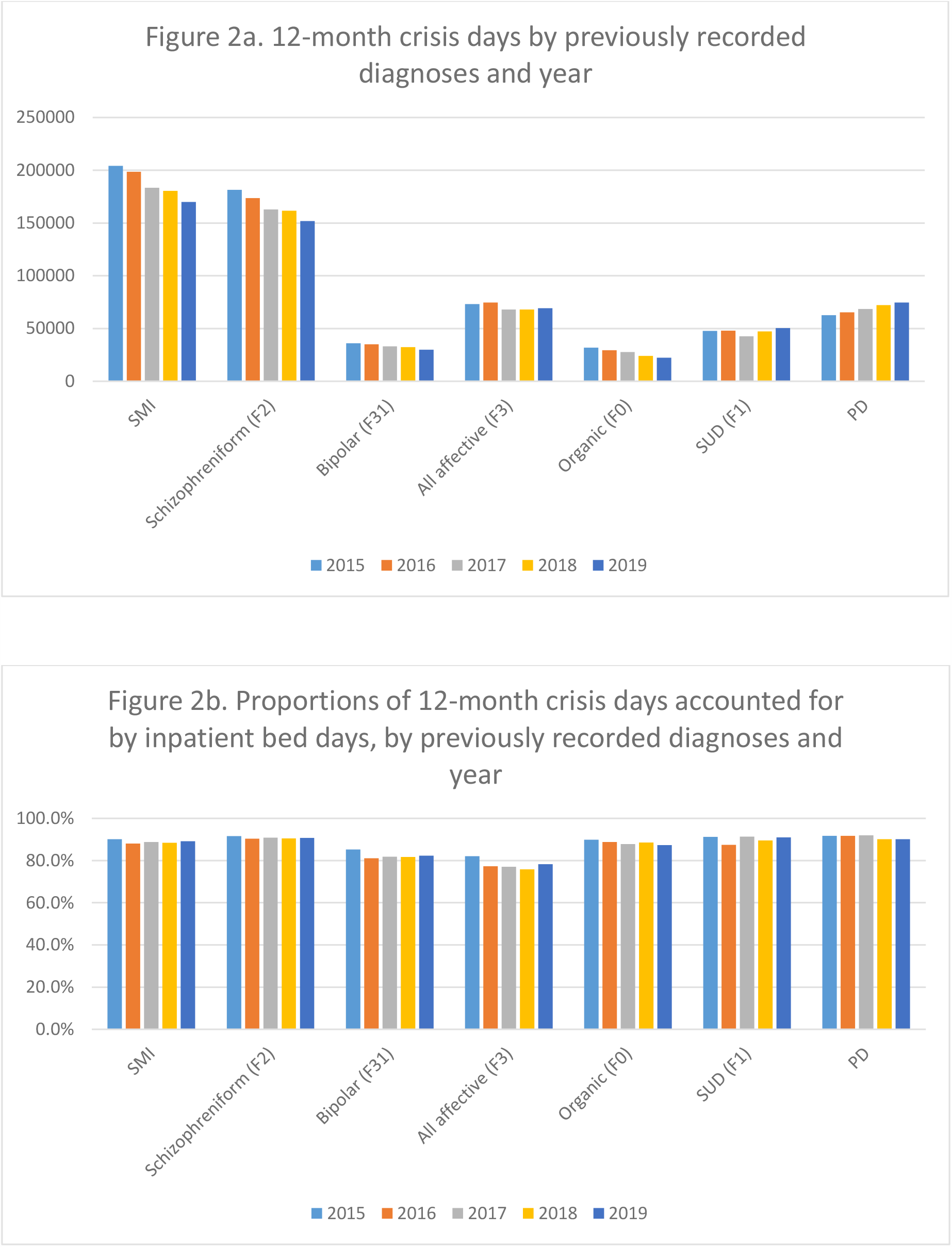
Crisis service use for the 12-month period following 16^th^ March in patients with current/recent mental healthcare at that point, displayed by year and presence or not of certain previously recorded diagnoses^1^.

Service contact changes for current/recent patients for the 31 day periods before and after 16^th^ March are compared by year in Table 2. In previous years, a consistent 3-6% reduction in inpatient person days was observed from periods before to after the index date, compared to the substantially larger 26% decrease observed in 2020. Face-to-face contacts showed a more heterogeneous 8-24% decrease over the same comparison period, compared to the 65% decrease in 2020, while virtual contacts had decreased by 3-22% in previous years compared to a 117% increase in 2020. Total contacts decreased by 21% in 2020, which was within the 8-23% range for previous years.

**Table 2.**
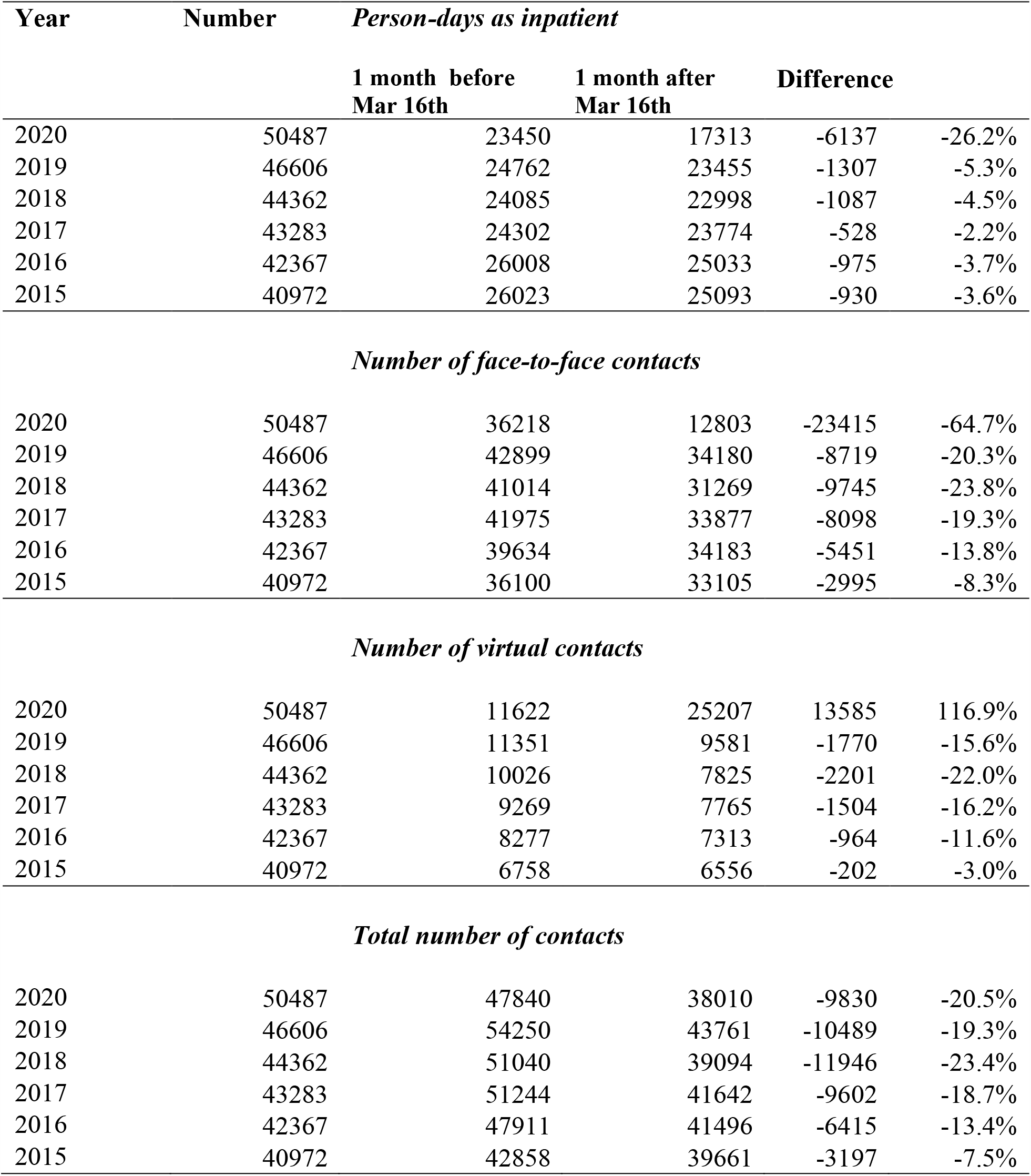
Service contact changes 31 days before and after March 16th in current/recent patients.^2^

| Year | Number | *Person-days as inpatient* |  |  |  |
| --- | --- | --- | --- | --- | --- |
|  |  | 1 month before<br>Mar 16th | 1 month after<br>Mar 16th | Difference |  |
| 2020 | 50487 | 23450 | 17313 | -6137 | -26.2% |
| 2019 | 46606 | 24762 | 23455 | -1307 | -5.3% |
| 2018 | 44362 | 24085 | 22998 | -1087 | -4.5% |
| 2017 | 43283 | 24302 | 23774 | -528 | -2.2% |
| 2016 | 42367 | 26008 | 25033 | -975 | -3.7% |
| 2015 | 40972 | 26023 | 25093 | -930 | -3.6% |

| *Number of face-to-face contacts* |  |  |  |  |  |
| --- | --- | --- | --- | --- | --- |
| 2020 | 50487 | 36218 | 12803 | -23415 | -64.7% |
| 2019 | 46606 | 42899 | 34180 | -8719 | -20.3% |
| 2018 | 44362 | 41014 | 31269 | -9745 | -23.8% |
| 2017 | 43283 | 41975 | 33877 | -8098 | -19.3% |
| 2016 | 42367 | 39634 | 34183 | -5451 | -13.8% |
| 2015 | 40972 | 36100 | 33105 | -2995 | -8.3% |

| *Number of virtual contacts* |  |  |  |  |  |
| --- | --- | --- | --- | --- | --- |
| 2020 | 50487 | 11622 | 25207 | 13585 | 116.9% |
| 2019 | 46606 | 11351 | 9581 | -1770 | -15.6% |
| 2018 | 44362 | 10026 | 7825 | -2201 | -22.0% |
| 2017 | 43283 | 9269 | 7765 | -1504 | -16.2% |
| 2016 | 42367 | 8277 | 7313 | -964 | -11.6% |
| 2015 | 40972 | 6758 | 6556 | -202 | -3.0% |

| *Total number of contacts* |  |  |  |  |  |
| --- | --- | --- | --- | --- | --- |
| 2020 | 50487 | 47840 | 38010 | -9830 | -20.5% |
| 2019 | 46606 | 54250 | 43761 | -10489 | -19.3% |
| 2018 | 44362 | 51040 | 39094 | -11946 | -23.4% |
| 2017 | 43283 | 51244 | 41642 | -9602 | -18.7% |
| 2016 | 42367 | 47911 | 41496 | -6415 | -13.4% |
| 2015 | 40972 | 42858 | 39661 | -3197 | -7.5% |

<sup>2</sup> Those with any active SLaM referral within 3 calendar months prior to that date.

Service contact changes are further displayed by previous diagnosis in Figures 3a-d. For inpatient days, most diagnostic groups showed substantial differences in changes between 2020 and previous years, apart from bipolar disorder where these were not marked. Patterns across years for changes in face-to-face, virtual, and total contacts did not differ substantially between diagnostic groups. Reductions in total contacts in 2020 were least marked for bipolar and schizophreniform disorders and most marked for organic disorders, but all of these changes were within the range for the previous 5 years.

**Figure 3.**
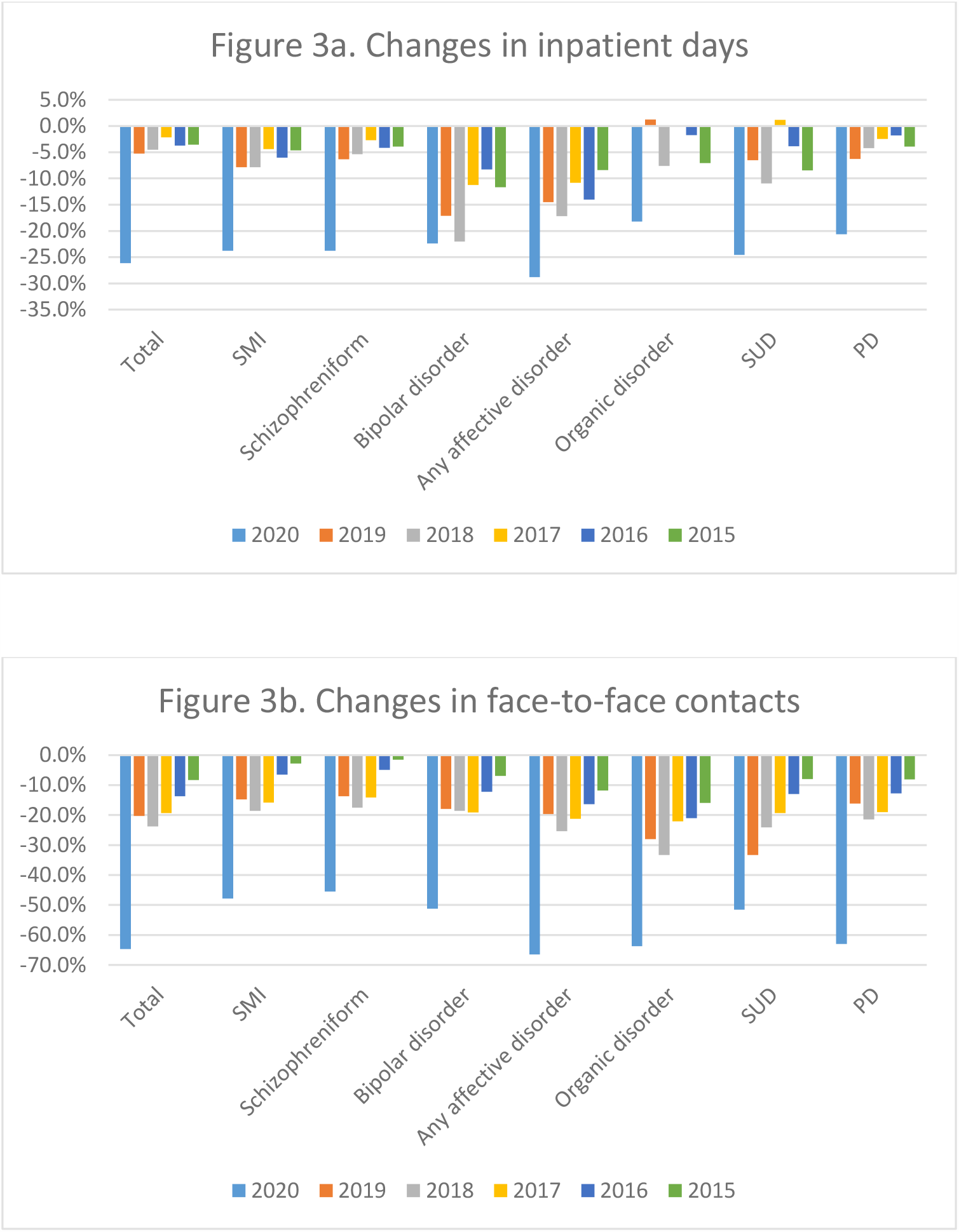
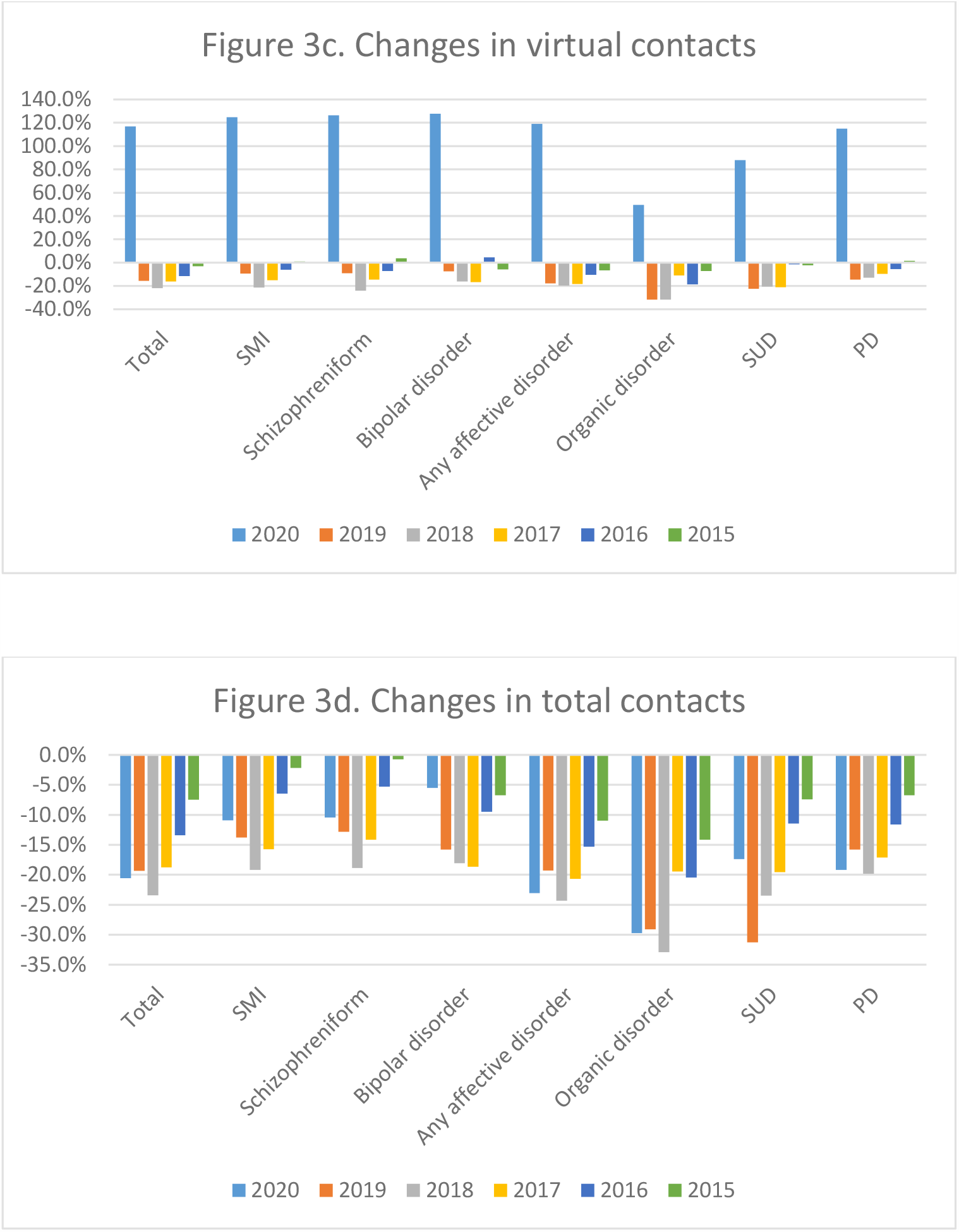
Proportional service contact changes^3^ from 31 days before to 31 days from/after March 16^th^ in current/recent patients, displayed by year and presence of previous recorded diagnosis^4^.

## Discussion

We report a series of descriptive analyses carried out at SLaM, carried out to inform the Trust on patients who might be most likely to require inpatient and other (HTT) crisis care in the 12 months following UK COVID-19 lockdown policy on 16^th^ March, based on findings from previous years, as well as to quantify levels of change in service delivery around that date for the group of current/recent service users most likely to be future users of crisis care. Key findings were as follows: i) 80-85% of crisis days were spent in inpatient care rather than under HTT; ii) around 75% of crisis days over a given 12 months were used by patients who were currently or recently under the care of the Trust at the commencement of that follow-up period; iii) highest numbers of crisis days were used by patients with a previously recorded schizophreniform disorder (ICD-10 F2x); iv) in recent/current patients, there had been substantial reductions in use of inpatient care in the 31 days after 16^th^ March 2020, substantially more than in previous years; v) changes in total contacts from non-inpatient teams did not differ substantially in 2020 compared to previous years, although there had been a marked change from face-to-face to virtual contacts – this held for all diagnostic groups investigated.

Considering limitations, it is important to bear in mind that the data are derived from a single site. Because complete data are being provided for that site with no hypothetical source population intended, calculation of confidence intervals was not felt to be appropriate for the descriptive data provided in this report; applicability to other mental healthcare providers cannot therefore be inferred and would need specific investigation, although we recommend the broad approach as a methodology worth considering. Profiles of services and catchment morbidity are also likely to vary between sites; for example, the very high predominance of people with psychotic disorders in those requiring inpatient and/or HTT input may well be related to SLaM’s particular inner urban catchment. Finally, the core task of predicting levels of future service activity, and the characteristics of those requiring highest-cost services, is clearly complex. On the one hand, there has been a marked consistency in the users of crisis services in past years; on the other hand, the COVID-19 pandemic has placed unique challenges on mental health services, those using them and those not yet using them but in need of them. Therefore it will be important to set up an adequate monitoring system to assess service use as close to real-time as possible and to identify any unexpected divergences from previous norms.

## Data Availability

Source data are available on request from the corresponding author.

## Funding

The research leading to these results has received support from the Medical Research Council Mental Health Data Pathfinder Award to King’s College London. RS and MB are part-funded by the National Institute for Health Research (NIHR) Biomedical Research Centre at the South London and Maudsley NHS Foundation Trust and King’s College London; RS is additionally part-funded by: i) a Medical Research Council (MRC) Mental Health Data Pathfinder Award to King’s College London; ii) an NIHR Senior Investigator Award; iii) the National Institute for Health Research (NIHR) Applied Research Collaboration South London (NIHR ARC South London) at King’s College Hospital NHS Foundation Trust. The views expressed are those of the authors and not necessarily those of the NIHR or the Department of Health and Social Care.

## Footnotes

1 SMI – serious mental illness; SUD – alcohol/substance use disorder; PD – personality disorder. Diagnoses based on any received up to 15^th^ March of the year in question.

2 Those with any active SLaM referral within 3 calendar months prior to that date.

3 Changes are expressed as percentages of the activity level in the 31 days prior to the 16^th^ March index date

4 SMI – serious mental illness; SUD – alcohol/substance use disorder; PD – personality disorder. Diagnoses based on any received up to 15^th^ March of the year in question.

## Notes

### Competing Interest Statement

RS declares research funding/support in the last 36 months from Janssen, GSK and Takeda.

### Author Declarations

CRIS has received approval as a data source for secondary analyses (Oxford Research Ethics Committee C, reference 18/SC/0372).

